# Generalised anxiety as a mediator linking ADHD symptoms and autistic traits to digital addictions in Australian emerging adults

**DOI:** 10.64898/2026.09.06.26362218

**Authors:** Matthew J. Giblett, Cara Jaklich, Brooke Andrew, Penelope A. Lind

## Abstract

**Background and aims:** Digital addiction is highly prevalent among emerging adults and is associated with significant detrimental health outcomes. Through the lens of the Interaction of Person-Affect-Cognition-Execution model, attention-deficit/hyperactivity disorder (ADHD) symptoms and autistic traits may increase vulnerability to digital addictions both directly through executive functioning deficits and indirectly through affective-cognitive processes such as anxiety. This study estimated whether internet, mobile phone, and video game addictions were represented by a common latent construct, estimated the associations of ADHD symptoms, autistic traits, generalised anxiety, and participant sex with these addictions, and estimated the extent to which anxiety mediated associations of ADHD symptoms and autistic traits with these addictions.

**Design:** Cross-sectional observational study.

**Setting:** Online survey conducted at Queensland University of Technology, Brisbane, Australia.

**Participants:** The analyses included 222 emerging adults aged 17–29 years (77.0% female) recruited between July and October 2015.

**Measurements:** The primary outcomes were the Internet Addiction Test (IAT), Mobile Phone Problem Use Scale (MPPUS), and Video Game Addiction Test (VAT). Predictors included the Autism Spectrum Quotient–10 (AQ-10), Adult ADHD Self-Report Scale v1.1 screener (ASRS-S), Generalised Anxiety Disorder-7 (GAD-7), and participant sex.

**Findings:** Internet, mobile phone, and video game addictions were not well represented by a single latent construct. GAD-7 was positively associated with AQ-10 (*B* = 0.20, 95% confidence interval [CI] [0.04, 0.37], *p* = .020) and ASRS-S (*B* = 0.54, CI [0.38, 0.70], *p* < .001). GAD-7 was positively associated with VAT (*B* = 0.63, CI [0.23, 1.00], *p* = .003), but neither IAT nor MPPUS. ASRS-S showed positive direct effects across IAT (*B* = 0.56, CI [0.08, 0.99], *p* = .024), MPPUS (*B* = 2.63, CI [1.06, 4.08], *p* < .001), and VAT (*B* = 0.41, CI [0.06, 0.78], *p* = .024), and a small-to-moderate positive indirect effect on VAT through GAD-7 (*B* = 0.35, CI [0.12, 0.60], *p* = .013). Sex did not moderate the associations between GAD-7 and digital addictions. Females reported higher MPPUS (*B* = 13.16, CI [6.17, 19.69], *p* < .001) and lower VAT (*B* = −5.22, CI [−7.00, −3.48], *p* < .001) scores than males.

**Conclusions:** Attention-deficit/hyperactivity disorder symptoms were associated with all three forms of digital addiction and showed an indirect association with video game addiction through generalised anxiety, consistent with the proposed mechanisms in the Interaction of Person-Affect-Cognition-Execution model.

## Introduction

Digital engagement is near-universal among emerging adults in Australia. Nearly all young adults use digital devices to access the internet, with mobile phones serving as the primary means of going online (Australian Communications and Media Authority, 2026). Over three-quarters of emerging adults report playing video games, often as a means of social connection (J. E. Brand & Jervis, 2021). Such widespread and intensive use of digital technologies has contributed to the emergence of digital addiction, comprising a range of problems arising from excessive engagement with digital media (Singh & Singh, 2019). However, the extent to which these problematic behaviours across different digital media reflect a single underlying construct remains unclear. Digital addiction is highly prevalent among emerging adults and university students globally (Fraiwan & Almomani, 2025; Liu et al., 2025; Salari et al., 2025; Wang & Abdullah, 2026) and is associated with significant and wide-ranging detrimental health outcomes (González-Bueso et al., 2018; Kalınkara & Talan, 2025; Kaya et al., 2025; Salman et al., 2025; Shiferaw et al., 2025).

The Interaction of Person-Affect-Cognition-Execution (I-PACE) model proposes that interactions between vulnerability factors and affective-cognitive processes contribute to the development and maintenance of addiction (M. Brand et al., 2016, 2019). Emerging adulthood is characterised by major life transitions and instability, including entry into higher education and the workforce (Fegert et al., 2025). University students may be particularly vulnerable to digital addiction due to both developmental factors and reliance on digital technologies for study (Tülübaş et al., 2023). Attention-deficit/hyperactivity disorder (ADHD) symptoms and autistic traits are relatively common among university students and are often co-occurring (Garcha & Smith, 2023; Nankoo et al., 2019). Higher levels of these traits are associated with greater depression, anxiety, and stress (Garcha & Smith, 2023; Nankoo et al., 2019; Waite et al., 2022). Consistent with the I-PACE model, ADHD symptoms and autistic traits may act as predisposing variables to addiction through impairments in executive functioning (Kofler et al., 2024; Southon, 2022; Townes et al., 2023). Importantly, neurodiverse traits and diagnoses are associated with both substance-related (Lyvers et al., 2024; Wilens et al., 2011) and behavioural addictions in adults (Fatséas et al., 2016; Grant & Chamberlain, 2021), including numerous forms of digital addiction (Cekic et al., 2024; Concerto et al., 2021; González-Bueso et al., 2018; Ho et al., 2014; Köse & Umut, 2024; Slack et al., 2025; Sulla et al., 2023). Additionally, individuals may respond to anxiety by using digital technologies to elevate their mood or avoid negative mood states, encouraging habitual use of digital technologies, whereby anxiety catalyses the development and perpetuation of digital addiction, which in turn perpetuates higher levels of anxiety (Kim et al., 2022). Therefore, higher levels of ADHD symptoms and autistic traits may increase vulnerability to digital addiction both directly, through executive functioning impairments, and indirectly via elevated anxiety.

This study employed structural equation modelling to estimate whether internet, mobile phone, and video game addictions were represented by a common latent construct, and to estimate the associations of ADHD symptoms, autistic traits, generalised anxiety, and participant sex with these digital addictions among university students. We hypothesised that the three digital addictions could be represented by a single latent factor. Additionally, we hypothesised that ADHD symptoms and autistic traits would positively predict anxiety, and that all three variables would show positive associations digital addiction. Given established sex differences in anxiety and digital behaviours, we also examined sex as a moderator of the association between anxiety and digital addiction to investigate whether this pathway operates differently across sexes. Finally, we hypothesised a partial mediation model in which anxiety mediates the effects of ADHD symptoms and autistic traits on digital addiction.

## Methods

### Participants

A total of 232 participants completed an anonymous online self-report questionnaire hosted by Qualtrics between July and October of 2015. Recruitment was via university flyers, online media appeals, and SONA Systems, where Queensland University of Technology (QUT) students enrolled through the SONA credit system received credit for their courses for participation. The questionnaire measured internet, gaming, and mobile phone/tablet use, as well as demographics, substance use, psychological health, social networks, and attitudes and motives around internet use and gaming. Participants’ sex was measured via self-report and included as a categorical variable (–1 = male, 1 = female). Participants gave informed consent through completing and submitting the questionnaire. Ten participants above the age of 29 were excluded from this study.

### Measures

#### Autism Spectrum Quotient

The adult version Autism Spectrum Quotient (AQ-10, Allison et al., 2012) is a 10-item measure of autistic traits in adults who do not have a learning disability. Participants rate each item on a 4-point Likert scale (1 = *definitely agree* to 4 = *definitely disagree*), with scores ranging from 10–40 and higher scores indicating greater trait expression. The suggested cutoff on this scale corresponds to a total score of 24, though this has not been validated. Although originally demonstrating good reliability, concerns have been raised regarding its psychometric properties in university samples (Cheung et al., 2023). Reliability was poor in this study (α = 0.52).

#### Adult ADHD Self-Report Scale Screener

The Adult ADHD Self-Report Scale v1.1 screener (ASRS-S, Kessler et al., 2005) is a 6-item measure of ADHD symptom frequency experienced over the previous six months. Participants rate each item on a 5-point Likert scale (0 = *never* to 4 = *very often*), with scores ranging from 0–24 and higher scores indicating greater symptom frequency. A total score of 0–9 indicates low levels of ADHD symptom severity, 10–13 mild to moderate, 14–17 high, and 18–24 very high, based on percentiles from a non-ADHD sample (Adler et al., 2019; Hegarty et al., 2024). The ASRS-S has demonstrated good reliability and validity across cross-cultural contexts (Lewczuk et al., 2024). Reliability was acceptable in this study (α = 0.75).

#### Generalised Anxiety Disorder

The Generalised Anxiety Disorder-7 (GAD-7, Spitzer et al., 2006) is a 7-item measure of generalised anxiety symptom severity over the prior two weeks in adults. Participants rate each item on a 4-point Likert scale (0 = *not at all* to 3 = *nearly every day*), with scores ranging from 0–21 and higher scores indicating greater anxiety severity. Total scores of 5–9 indicate mild levels of anxiety, 10–14 moderate, and 15–21 severe. The GAD-7 demonstrated excellent internal consistency and good construct validity in university students (Byrd-Bredbenner et al., 2021; Dhira et al., 2021). Reliability was excellent in this study (α = 0.92).

#### Internet Addiction Test

The Internet Addiction Test (IAT, Young, 1998) is a 20-item measure of compulsive internet use severity over the prior 12 months in adults and adolescents. Participants rate each item on a 5-point Likert scale (0 = *rarely or never* to 4 = *always*), with scores ranging from 0–80 and higher scores indicating greater compulsive internet use severity. As our scoring differs slightly to the standard 6-point Likert scale, which includes a “does not apply” option, we offset the proposed cutoffs proportionally, with scores 0–24 indicating normal internet use, 25–39 mild addiction, 40–63 moderate, and 64–80 severe, however there is limited evidence for these thresholds (Sayili et al., 2025). The IAT demonstrated excellent reliability and good validity in university students (Coelho et al., 2025; Moon et al., 2018; Wei et al., 2025). Reliability was good in this study (α = 0.88).

#### Mobile Phone Problem Use Scale

The Mobile Phone Problem Use Scale (MPPUS, Bianchi & Phillips, 2005) is a 27-item measure of problematic mobile phone use severity over the prior 12 months in adults. Participants rate each item on a 10-point Likert scale (1 = *not at all true* to 10 = *extremely true*), with scores ranging from 27– 270 and higher scores indicating greater problematic mobile phone use severity. Consistent with prior research, we dropped item 4 (“All my friends own a mobile phone”) due to a prominent ceiling effect. Total scores of 26–32 indicate *Casual Users*, 33–96 *Habitual or Regular Users*, 97–138 *At Risk Users*, and 139–260 *Problematic Users* (de-Sola et al., 2017). This 26-item version demonstrated high internal consistency and good construct validity in Australian adults (Oviedo-Trespalacios et al., 2019). Reliability was excellent in this study (α = 0.94).

#### Video Game Addiction Test

The Video Game Addiction Test (VAT, Van Rooij et al., 2012) is a 14-item measure of problematic video game use severity over the prior 12 months. Participants rate each item on a 5-point Likert scale (0 = *never* to 4 = *very often*), with scores ranging from 0–56 and higher scores indicating greater problematic video game use severity. The VAT demonstrated excellent reliability and good construct validity in a sample of secondary school students from the Netherlands (Van Rooij et al., 2012). Reliability was excellent in this study (α = 0.95).

### Design and data analysis

This study uses a cross-sectional correlational design. The data analysis plan was preregistered with Open Science Framework on 22 April 2026, prior to data analysis (https://osf.io/swfuj). There were no deviations from this protocol. Analyses were performed using R (version 4.5.2, R Core Team, 2025) with the *lavaan* package (version 0.6-21, Rosseel, 2012; Rosseel et al., 2025) for confirmatory factor analysis (CFA) and path analysis. Prior to main analyses, we examined patterns of missing data and assessed assumptions including normality, multicollinearity, linearity, and outliers (see the Supplementary Material for details). Supplementary Tables S1–S4 and Supplementary Figures S1–S2 present some of the results of these analyses. Models were estimated using maximum likelihood (ML), with standard errors and confidence intervals obtained via nonparametric bootstrapping (5,000 resamples). Missing data were handled using full information maximum likelihood (FIML, Enders & Bandalos, 2001), which uses all available data under the assumption of missing at random (MAR). Exogenous variables were treated as random so that their variances and covariances were estimated and missing data on these variables could be handled using FIML. Model fit was evaluated using χ^2^, Comparative Fit Index (CFI), Tucker-Lewis Index (TLI), Root Mean Square Error of Approximation (RMSEA), and Standardized Root Mean Square Residual (SRMR).

A CFA was specified to model digital addiction as a latent construct, indicated by IAT, MPPUS, and VAT. The model was identified using marker-variable scaling by fixing the loading of one indicator to 1. Residual variances of the observed indicators were assumed to be uncorrelated. The structural model, specified following evaluation of the measurement model, included AQ-10, ASRS-S, and sex as exogenous predictors of GAD-7 and digital addiction; GAD-7 was included as an endogenous predictor of digital addiction. AQ-10 and ASRS-S were modelled as correlated exogenous variables, with their covariance freely estimated. Sex was included as a moderator of the association between generalised anxiety and digital addiction via an observed interaction term (Sex × GAD-7). GAD-7 was mean-centred to reduce multicollinearity with the interaction term. All tests used a two-tailed significance level of α = .05.

## Results

Table 1 displays descriptive statistics and sociodemographic characteristics of 222 participants and Table 2 presents correlations among study variables. The majority of participants were current QUT School of Psychology students at the time of the survey and born in Australia. Most participants were unemployed with a highest completed education of senior high school. Across measures, average scores generally fell within mild or subclinical ranges. GAD-7 scores were positively skewed with a moderate floor effect (see Supplementary Figure S1C), reflecting overall mild levels of generalised anxiety symptom severity. Average ASRS-S scores indicated mild to moderate levels of ADHD symptoms and average AQ-10 scores were below the cutoff consistent with autism. Average IAT scores indicated overall normal internet usage and average MPPUS scores suggested overall at risk of problematic mobile phone use. VAT scores were positively skewed with a pronounced floor effect (see Supplementary Figure S1F).

**Table 1.** Descriptive Statistics and Sociodemographic Characteristics of Participants.

| Characteristic | Sex |  | Total (*N* = 222) |
| --- | --- | --- | --- |
|  | Female (*n* = 171) | Male (*n* = 51) |  |
| Age at survey, years |  |  |  |
| Mean (*SD*) | 19.8 (2.7) | 20.7 (3.5) | 20.0 (2.9) |
| Min–Max | 17.0–29.0 | 17.0–29.0 | 17.0–29.0 |
| Missing, *n* (%) | 1 (0.6) | 1 (2.0) | 2 (0.9) |
| Country of birth, *n* (%) |  |  |  |
| Australia | 141 (82.5) | 47 (92.2) | 188 (84.7) |
| New Zealand | 6 (3.5) | 1 (2.0) | 7 (3.2) |
| USA | 3 (1.8) | 0 (0.0) | 3 (1.4) |
| England, Ireland, Scotland or Wales | 5 (2.9) | 0 (0.0) | 5 (2.3) |
| India | 2 (1.2) | 1 (2.0) | 3 (1.4) |
| Other | 14 (8.2) | 2 (3.9) | 16 (7.2) |
|  | Female (*n* = 171) | Male (*n* = 51) |  |
| QUT School of Psychology student at survey, *n* (%) |  |  |  |
| No | 29 (17.2) | 17 (34.7) | 46 (21.1) |
| Yes | 140 (82.8) | 32 (65.3) | 172 (78.9) |
| Missing, *n* (%) | 2 (1.2) | 2 (3.9) | 4 (1.8) |
| Highest level of education, *n* (%) |  |  |  |
| Certificate or Diploma | 20 (11.7) | 12 (23.5) | 32 (14.4) |
| Senior High School | 128 (74.9) | 33 (64.7) | 161 (72.5) |
| University degree (any level) | 23 (13.5) | 6 (11.8) | 29 (13.1) |
| Employment, *n* (%) |  |  |  |
| Employed | 79 (46.5) | 22 (43.1) | 101 (45.7) |
| Not employed | 91 (53.5) | 29 (56.9) | 120 (54.3) |
| Missing, *n* (%) | 1 (0.6) | 0 (0) | 1 (0.5) |
| Income, *n* (%) |  |  |  |
| ≤ \$39,999 | 23 (13.5) | 10 (19.6) | 33 (14.9) |
| ≥ \$40,000 | 52 (30.4) | 17 (33.3) | 69 (31.1) |
| Income unknown / not disclosed | 96 (56.1) | 24 (47.1) | 120 (54.1) |
| AQ-10 |  |  |  |
| Mean (*SD*) | 21.1 (3.6) | 20.7 (3.5) | 21.0 (3.5) |
| Min–Max | 14.0–34.0 | 13.0–29.0 | 13.0–34.0 |
| Missing, *n* (%) | 24 (14) | 12 (24) | 36 (16) |
| ASRS-S |  |  |  |
| Mean (*SD*) | 9.4 (4.3) | 9.5 (4.2) | 9.4 (4.2) |
| Min–Max | 0.0–24.0 | 1.0–18.0 | 0.0–24.0 |
| Missing, *n* (%) | 24 (14) | 12 (24) | 36 (16) |
| GAD-7 |  |  |  |
| Mean (*SD*) | 6.2 (5.3) | 4.7 (4.6) | 5.9 (5.2) |
| Min–Max | 0.0–21.0 | 0.0–16.0 | 0.0–21.0 |
| Missing, *n* (%) | 24 (14) | 12 (24) | 36 (16) |
| IAT |  |  |  |
| Mean (*SD*) | 16.8 (9.7) | 18.7 (13.0) | 17.2 (10.5) |
| Min–Max | 1.0–41.0 | 2.0–59.0 | 1.0–59.0 |
| Missing, *n* (%) | 17 (9.9) | 6 (12) | 23 (10) |
| MPPUS |  |  |  |
| Mean (*SD*) | 109.4 (40.3) | 80.8 (40.7) | 103.5 (41.9) |
| Min–Max | 28.0–210.0 | 26.0–196.0 | 26.0–210.0 |
| Missing, *n* (%) | 28 (16) | 14 (27) | 42 (19) |
| VAT |  |  |  |
| Mean (*SD*) | 7.1 (9.6) | 16.2 (11.7) | 9.2 (10.8) |
|  | Female (*n* = 171) | Male (*n* = 51) |  |
| Min–Max | 0.0–42.0 | 0.0–53.0 | 0.0–53.0 |
| Missing, *n* (%) | 41 (24) | 11 (22) | 52 (23) |

*Note.* QUT = Queensland University of Technology; AQ-10 = Autism Spectrum Quotient-10;
ASRS-S = Adult ADHD Self-Report Scale v1.1 screener; GAD-7 = Generalised Anxiety
Disorder-7; IAT = Internet Addiction Test; MPPUS = Mobile Phone Problem Use Scale;
VAT = Video Game Addiction Test.

**Table 2.**
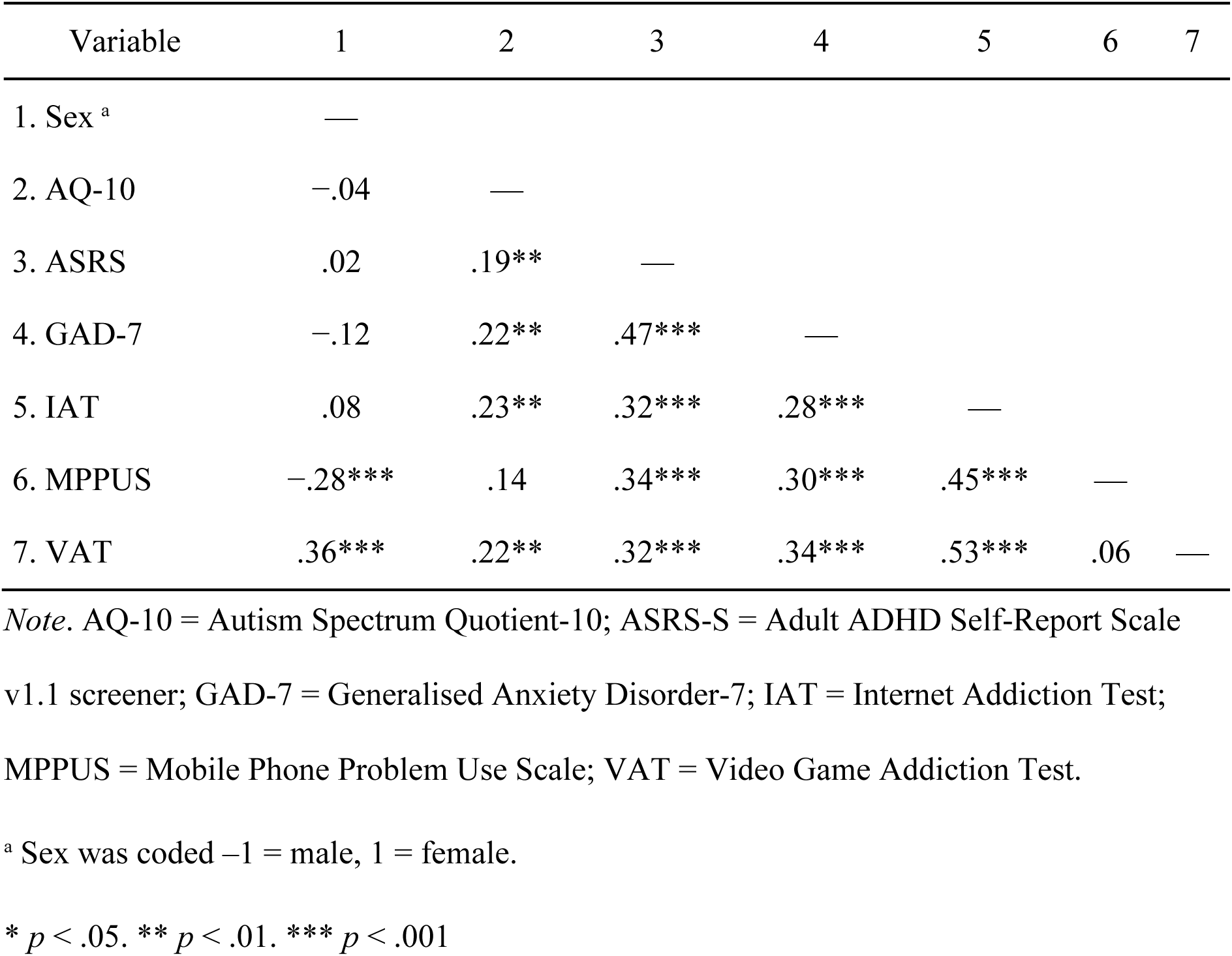
Pearson r Correlations Among Study Variables.

The global model fit of the CFA to assess whether IAT, MPPUS, and VAT loaded onto a single latent digital addiction factor could not be evaluated because a single-factor model with three indicators is just-identified, resulting in zero degrees of freedom. The estimated parameter of IAT was inadmissible (a Heywood case) with negative residual variance (−137.0) and a standardised estimate greater than one (β = 1.50), indicating model misspecification and suggesting that the latent factor does not adequately account for the covariance among the indicators. Loadings for MPPUS (*B* = 0.82, 95% CI [−0.62, 2.25], *p* = .264) and VAT (*B* = 0.24, CI [−0.18, 0.65], *p* = .258) were small and non-significant. Inspection of the correlations among the indicators (see Table 2) suggested that associations between the measures were inconsistent. Consistent with the preregistered analysis plan, we did not proceed with the latent variable structural model. Instead, we retained IAT, MPPUS, and VAT as separate outcomes in three subsequent path analyses. As a result of the additional tests, we controlled the false discovery rate (FDR) using the Benjamini–Hochberg procedure (Benjamini & Hochberg, 1995) applied separately within each set of parallel paths across the three models (e.g., across the GAD-7 → IAT, MPPUS, VAT paths).

Model fit was poor across all three models, as indicated by significant χ^2^ tests and suboptimal fit indices (see Supplementary Table S5). The Sex × GAD-7 interaction was non-significant across the models, providing no support for our hypothesis that sex moderates the relationship between generalised anxiety and digital addiction. See Supplementary Table S6 for all path parameters. Given the consistently poor model fit and non-significant moderation effects, we tested a more parsimonious mediation model excluding the interaction term. As shown in Table 3, all three final models demonstrated excellent fit to the data. Path parameters for these models are reported in Table 4 and diagrams with standardised coefficients shown in Figure 1. The proportion of variance explained (*R*^2^) for GAD-7 was 0.26, 0.16 for IAT, 0.21 for MPPUS, and 0.34 for VAT.

**Figure 1.**
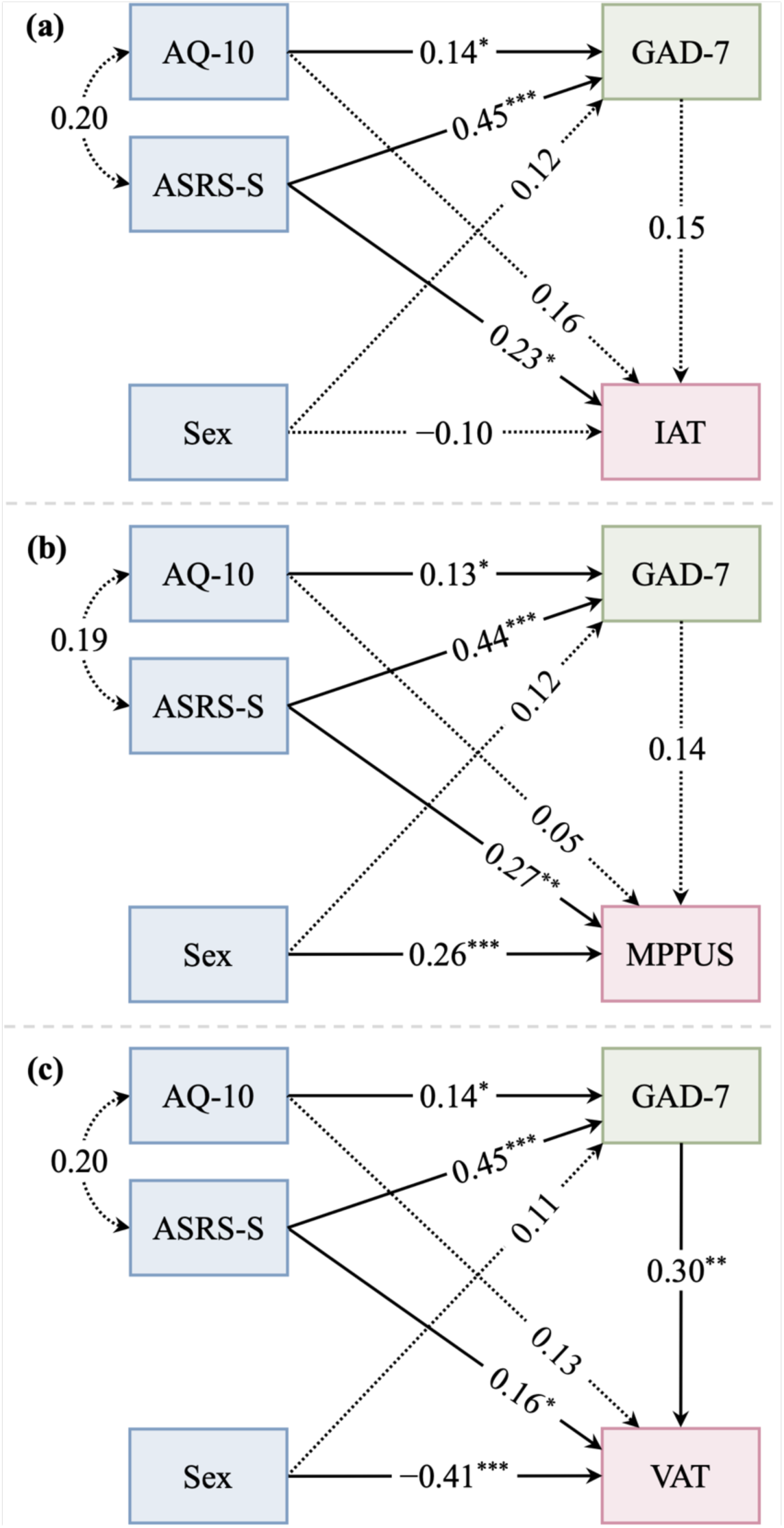
Diagrams of Path Analyses Examining Associations among AQ-10, ASRS-S, Participant Sex, GAD-7, and Digital Addictions. *Note.* Coefficients are standardised linear regression coefficients. Significance levels are Benjamini–Hochberg false discovery rate (FDR) adjusted. Blue represents exogenous predictors, green represents the endogenous mediator, and red represents the endogenous outcome. Solid paths represent significant coefficients, and dotted paths represent non-significant coefficients. (a) Path analysis of Autism Spectrum Quotient-10 (AQ-10), Adult ADHD Self-Report Scale v1.1 screener (ASRS-S), and participant sex (coded –1 = male, 1 = female) predicting Generalised Anxiety Disorder-7 (GAD-7) and Internet Addiction Test (IAT), and GAD-7 predicting IAT; (b) path analysis of AQ-10, ASRS-S, and sex predicting GAD-7 and Mobile Phone Problem Use Scale (MPPUS), and GAD-7 predicting MPPUS; (c) path analysis of AQ-10, ASRS-S, and sex predicting GAD-7 and Video Game Addiction Test (VAT), and GAD-7 predicting VAT. * *p* < .05. ** *p* < .01. *** *p* < .001

**Table 3.** Fit Indices of Path Analysis Models Examining Associations among AQ-10, ASRS-S, Participant Sex, GAD-7, and Digital Addiction Outcomes (IAT, MPPUS, and VAT)

| Model | Chi-Square Goodness of Fit |  |  | Incremental Fit |  | Absolute Fit |  |  |
| --- | --- | --- | --- | --- | --- | --- | --- | --- |
| | $\chi^2$ | *df* | *p* | CFI | TLI | RMSEA | 95% CI | SRMR |
| IAT | 0.41 | 2 | .814 | 1.00 | 1.10 | 0.00 | [0.00, 0.08] | 0.01 |
| MPPUS | 0.42 | 2 | .811 | 1.00 | 1.08 | 0.00 | [0.00, 0.08] | 0.01 |
| VAT | 0.42 | 2 | .809 | 1.00 | 1.07 | 0.00 | [0.00, 0.08] | 0.01 |

*Note.* CI = confidence interval; CFI = Comparative Fit Index; TLI = Tucker-Lewis Index; RMSEA = Root Mean Square Error of Approximation; SRMR = Standardized Root Mean Square Residual; IAT = Internet Addiction Test; MPPUS = Mobile Phone Problem Use Scale; VAT = Video Game Addiction Test.

**Table 4.** Parameter Estimates of Path Analyses Examining Associations among AQ-10, ASRS-S, Participant Sex, GAD-7, and Digital Addictions.

| Parameter | IAT |  |  |  |  | MPPUS |  |  |  |  | VAT |  |  |  |  |
| --- | --- | --- | --- | --- | --- | --- | --- | --- | --- | --- | --- | --- | --- | --- | --- |
| | *B* | 95% CI <sup>a</sup> | *SE* <sup>a</sup> | $\beta$ | *p* FDR <sup>b</sup> | *B* | 95% CI <sup>a</sup> | *SE* <sup>a</sup> | $\beta$ | *p* FDR <sup>b</sup> | *B* | 95% CI <sup>a</sup> | *SE* <sup>a</sup> | $\beta$ | *p* FDR <sup>b</sup> |
| **Covariances** |  |  |  |  |  |  |  |  |  |  |  |  |  |  |  |
| AQ-10 ↔ ASRS-S | 2.96 | [0.47, 5.65] | 1.35 | 0.20 | .055 | 2.88 | [0.41, 5.54] | 1.33 | 0.19 | .055 | 2.95 | [0.47, 5.64] | 1.35 | 0.20 | .055 |
| **Direct effects on GAD-7** |  |  |  |  |  |  |  |  |  |  |  |  |  |  |  |
| AQ-10 | 0.20 | [0.04, 0.37] | 0.08 | 0.14 | .020* | 0.19 | [0.03, 0.36] | 0.08 | 0.13 | .020* | 0.20 | [0.04, 0.37] | 0.08 | 0.14 | .020* |
| ASRS-S | 0.54 | [0.38, 0.70] | 0.08 | 0.45 | < .001*** | 0.54 | [0.38, 0.70] | 0.08 | 0.44 | < .001*** | 0.54 | [0.38, 0.71] | 0.08 | 0.45 | < .001*** |
| Sex <sup>c</sup> | 0.72 | [0.01, 1.42] | 0.36 | 0.12 | .057 | 0.74 | [0.03, 1.46] | 0.36 | 0.12 | .056 | 0.69 | [-0.03, 1.40] | 0.36 | 0.11 | .063 |
| **Direct effects on digital addiction** |  |  |  |  |  |  |  |  |  |  |  |  |  |  |  |
| GAD-7 | 0.31 | [0.00, 0.65] | 0.17 | 0.15 | .091 | 1.12 | [-0.20, 2.44] | 0.67 | 0.14 | .091 | 0.63 | [0.23, 1.00] | 0.19 | 0.30 | .003** |
| AQ-10 | 0.48 | [0.07, 0.94] | 0.22 | 0.16 | .093 | 0.56 | [-1.06, 2.31] | 0.86 | 0.05 | .512 | 0.39 | [-0.07, 0.84] | 0.22 | 0.13 | .125 |
| ASRS-S | 0.56 | [0.08, 0.99] | 0.23 | 0.23 | .024* | 2.63 | [1.06, 4.08] | 0.76 | 0.27 | .002** | 0.41 | [0.06, 0.78] | 0.18 | 0.16 | .024* |
| Sex <sup>c</sup> | -1.19 | [-3.17, 0.65] | 0.97 | -0.10 | .220 | 13.16 | [6.17, 19.69] | 3.44 | 0.26 | < .001*** | -5.22 | [-7.00, -3.48] | 0.89 | -0.41 | < .001*** |
| **Indirect effects on digital addiction through GAD-7** |  |  |  |  |  |  |  |  |  |  |  |  |  |  |  |
| AQ-10 | 0.06 | [0.00, 0.17] | 0.05 | 0.02 | .189 | 0.22 | [-0.04, 0.60] | 0.17 | 0.02 | .189 | 0.13 | [0.02, 0.29] | 0.07 | 0.04 | .189 |
| ASRS-S | 0.17 | [0.00, 0.37] | 0.09 | 0.07 | .106 | 0.61 | [-0.10, 1.41] | 0.38 | 0.06 | .112 | 0.35 | [0.12, 0.60] | 0.12 | 0.14 | .013* |
| | *B* | 95% CI <sup>a</sup> | *SE* <sup>a</sup> | $\beta$ | *p* FDR <sup>b</sup> | *B* | 95% CI <sup>a</sup> | *SE* <sup>a</sup> | $\beta$ | *p* FDR <sup>b</sup> | *B* | 95% CI <sup>a</sup> | *SE* <sup>a</sup> | $\beta$ | *p* FDR <sup>b</sup> |
| **Total effects on digital addiction** |  |  |  |  |  |  |  |  |  |  |  |  |  |  |  |
| AQ-10 | 0.54 | [0.12, 1.01] | 0.23 | 0.18 | .037* | 0.78 | [-0.84, 2.52] | 0.84 | 0.07 | .354 | 0.51 | [0.04, 0.97] | 0.23 | 0.17 | .037* |
| ASRS-S | 0.72 | [0.28, 1.14] | 0.22 | 0.29 | <.001*** | 3.23 | [1.76, 4.55] | 0.69 | 0.33 | <.001*** | 0.76 | [0.38, 1.15] | 0.19 | 0.30 | <.001*** |

*Note.* CI = confidence interval; FDR = false discovery rate; AQ-10 = Autism Spectrum Quotient-10; ASRS-S = Adult ADHD Self-Report Scale v1.1 screener; GAD-7 = Generalised Anxiety Disorder-7; IAT = Internet Addiction Test; MPPUS = Mobile Phone Problem Use Scale; VAT = Video Game Addiction Test.
<sup>a</sup> *SEs* and CIs were obtained via nonparametric bootstrapping (5,000 resamples).
<sup>b</sup> *p*-values are Benjamini–Hochberg FDR adjusted across each model.
<sup>c</sup> Sex was coded –1 = male, 1 = female.
\* $p < .05$ . \*\* $p < .01$ . \*\*\* $p < .001$

ASRS-S and AQ-10 were significantly positively associated with GAD-7, whereas sex was not significantly associated with GAD-7. GAD-7 was significantly positively associated with VAT but was not significantly associated with IAT or MPPUS. AQ-10 showed no significant direct effects on any of the digital addiction measures. In contrast, ASRS-S demonstrated significant positive direct effects on VAT, IAT, and MPPUS. We did not observe any significant indirect effects of AQ-10 on digital addiction outcomes through GAD-7. However, we did find a significant small-to-moderate positive indirect effect of ASRS-S via GAD-7 on VAT, indicating mediation for video game addiction only. No significant indirect effects of ASRS-S through GAD-7 were found for IAT or MPPUS. AQ-10 and ASRS-S did not significantly covary after FDR correction. No significant sex differences were observed for IAT. Females reported significantly higher MPPUS scores, whereas males reported significantly higher VAT scores.

## Discussion

This study estimated the relationships between ADHD symptoms, autistic traits, generalised anxiety, and internet, mobile phone, and video gaming addictions. We also estimated whether these behaviours could be conceptualised as related and overlapping forms of problematic digital engagement. Overall, the findings provided no support for an overlapping digital addiction construct and partial support for our proposed path analysis model, highlighting ADHD symptoms as a robust predictor of digital addictions, while offering more limited and domain-specific support for the roles of generalised anxiety and sex. GAD-7 scores in our sample were comparable to those reported in Australian community samples (Stocker et al., 2021). The pronounced floor effect in VAT scores was also consistent with prior research in non-clinical samples showing that item-level scores largely fall between 0 and 1 out of 4 (Van Rooij et al., 2017).

The CFA of IAT, MPPUS, and VAT found no evidence supporting a latent digital addiction construct, indicating that these addictive behaviours function as unique and separate forms of problematic digital engagement. However, it is possible that overuse of one digital media may confer risk for overuse of others. For example, people who overuse the internet or video gaming may do so on their phone, potentially developing a combination of all three addictions.

The I-PACE model (M. Brand et al., 2016, 2019) suggests that individuals experiencing high levels of anxiety may seek to resolve negative affect by using digital technologies, such as video games (Kim et al., 2022). Our findings provide limited support for the hypothesis that generalised anxiety serves as a broad predictor of digital addiction. Although GAD-7 was positively associated with video game addiction, it was not significantly related to internet addiction or problematic mobile phone use. This suggests that generalised anxiety may be more relevant to certain forms of digital addiction than others. This finding contrasts research on the fear of missing out, where students may use smartphones and social media as a relief from the anxiety of missing out on future social experiences (Elhai et al., 2025).

AQ-10 was positively associated with GAD-7, supporting prior links between autistic traits and anxiety (Garcha & Smith, 2023) and indicating that emerging adults with higher levels of autistic traits may be at greater risk of experiencing anxiety. However, our results did not support the hypothesised role of autistic traits as a predisposing factor for digital addiction. AQ-10 was not directly associated with any of the digital addiction measures and did not exert indirect effects through generalized anxiety, contrasting prior research showing positive effects of autistic traits on internet addiction (Köse & Umut, 2024), problematic mobile phone use (Zhou et al., 2024), and video game addiction (Carpita et al., 2026; Concerto et al., 2021; Slack et al., 2025). Nevertheless, the AQ-10 showed poor reliability which may help explain our lack of expected effects and also supports concerns about its psychometric properties (Cheung et al., 2023). Therefore, these results should be interpreted with caution, and future studies should replicate these findings using different measures of autistic traits.

Unlike the AQ-10, the ASRS-S emerged as a consistent predictor of digital addiction, with significant positive direct effects across internet addiction, problematic mobile phone use, and video game addiction. This pattern supports the hypothesised role of ADHD symptoms on digital addiction and is consistent with the I-PACE model, suggesting that executive functioning impairments associated with ADHD, such as poor inhibitory control and increased reward sensitivity (Senkowski et al., 2024; von Rhein et al., 2015), may broadly increase susceptibility to problematic digital technology use. These findings are also consistent with previous studies linking ADHD traits to heightened risk for both substance-related and behavioural addictions (Concerto et al., 2021; Fatséas et al., 2016; Lyvers et al., 2024). While the AQ-10 includes two items related to executive function (attention-switching and multi-tasking), it is not a measure of executive functioning. This may explain why we observed more effects of the ASRS-S on digital addiction and not the AQ-10. Mediation analyses revealed a small-to-moderate indirect effect of ASRS-S on VAT via GAD-7, but no comparable effects for IAT or MPPUS. This suggests that generalised anxiety may act as a pathway linking ADHD symptoms to video gaming behaviours specifically, rather than a general mediator across all forms of digital addiction.

Sex differences showed a domain-specific pattern, with females reporting higher problematic mobile phone use and males higher video game addiction. No sex differences emerged for internet addiction or generalised anxiety, and sex did not moderate the association between anxiety and digital addiction outcomes. These findings suggest that while males and females may differ in the types of digital activities they engage in, the effects, or lack thereof, of generalised anxiety on digital addictions operates similarly across sexes.

Our findings support prior research suggesting the importance of identifying university students with high risk of ADHD to reduce or prevent negative mental health outcomes, such as anxiety and digital addictions (Nankoo et al., 2019). Additionally, our results reinforce the importance of considering the role that mental health conditions, such as anxiety, ADHD, and autism, play in the expression of digital addiction.

### Limitations

We cannot infer causality as this is a cross-sectional study. All measures were self-report questionnaires that were freely available for research use. Given that our convenience sample consists primarily of QUT School of Psychology students, our results are at best generalisable to emerging adult Australian university students rather than the broader Australian community (Andrade, 2021). Future studies could work to replicate our findings in broader samples to improve its external validity. Additionally, the findings may not reflect current digital media use due to significant changes in digital technologies over the past decade, such as the rise of large language models (LLMs) which also foster addictive behaviours (Yankouskaya et al., 2025). Future research could also investigate the effects of mental health issues on LLM overuse or addiction.

## Statements & Declarations

## Acknowledgments

This research was conducted on the Country of the Yuggera and Turrbal peoples. We acknowledge the Yuggera and Turrbal peoples as Traditional Owners and custodians of these lands. We pay our respects to their Ancestors and descendants, who continue to have cultural and spiritual connections to Country. We recognise their valuable contributions to Australian and global society. We thank Lee Jones and the QIMR Berghofer Statistics Unit for assisting with the statistical design and the participants for their time and support.

## Author Contributions

Author roles were classified using the Contributor Role Taxonomy (CRediT; https://credit.niso.org/) as follows: Matthew J. Giblett: conceptualisation, data curation, formal analysis, investigation, visualisation, writing – original draft; Cara Jaklich: conceptualisation, investigation, methodology, project administration, writing – review & editing; Brooke Andrew: conceptualisation, methodology, project administration, supervision, writing – review & editing; Penelope A. Lind: conceptualisation, methodology, project administration, supervision, writing – review & editing.

## Consent to Participate

Submission of the completed online questionnaire was accepted as an indication of participant consent.

## Data Availability

Anonymised survey data may be shared for collaborative projects, subject to a data transfer agreement and governance and ethics approval.

## Ethics Approval

Ethical approval was obtained prior to data collection from the University Human Research Ethics Committee of QUT (approval number: 1500000433). Ethical approval for all aspects of this work was obtained from QIMR Berghofer Human Research Ethics Committee (P1559).

## References

Adler, L. A., Faraone, S. V., Sarocco, P., Atkins, N., & Khachatryan, A. (2019). Establishing US norms for the Adult ADHD Self-Report Scale (ASRS-v1.1) and characterising symptom burden among adults with self-reported ADHD. International Journal of Clinical Practice, 73(1), Article e13260. 10.1111/ijcp.13260

Allison, C., Auyeung, B., & Baron-Cohen, S. (2012). Toward brief “red flags” for autism screening: The short Autism Spectrum Quotient and the short quantitative checklist in 1,000 cases and 3,000 controls. Journal of the American Academy of Child & Adolescent Psychiatry, 51(2), 202–212. 10.1016/j.jaac.2011.11.003

Andrade, C. (2021). The inconvenient truth about convenience and purposive samples. Indian Journal of Psychological Medicine, 43(1), 86–88. 10.1177/0253717620977000

Australian Communications and Media Authority. (2026). Communications and media in Australia: How we use the internet. https://www.acma.gov.au/communications-and-media-australia

Benjamini, Y., & Hochberg, Y. (1995). Controlling the false discovery rate: A practical and powerful approach to multiple testing. Journal of the Royal Statistical Society Series B: Statistical Methodology, 57(1), 289–300. 10.1111/j.2517-6161.1995.tb02031.x

Bianchi, A., & Phillips, J. G. (2005). Psychological predictors of problem mobile phone use. Cyberpsychology & Behavior: The Impact of the Internet, Multimedia and Virtual Reality on Behavior and Society, 8(1), 39–51. 10.1089/cpb.2005.8.39

Brand, J. E., & Jervis, J. (2021). Digital Australia 2022. Interactive Games and Entertainment Association. https://igea.net/wp-content/uploads/2021/10/DA22-Report-FINAL-19-10-21.pdf

Brand, M., Wegmann, E., Stark, R., Müller, A., Wölfling, K., Robbins, T. W., & Potenza, M. N. (2019). The Interaction of Person-Affect-Cognition-Execution (I-PACE) model for addictive behaviors: Update, generalization to addictive behaviors beyond internet-use disorders, and specification of the process character of addictive behaviors. Neuroscience & Biobehavioral Reviews, 104, 1–10. 10.1016/j.neubiorev.2019.06.032

Brand, M., Young, K. S., Laier, C., Wölfling, K., & Potenza, M. N. (2016). Integrating psychological and neurobiological considerations regarding the development and maintenance of specific Internet-use disorders: An Interaction of Person-Affect-Cognition-Execution (I-PACE) model. Neuroscience & Biobehavioral Reviews, 71, 252–266. 10.1016/j.neubiorev.2016.08.033

Byrd-Bredbenner, C., Eck, K., & Quick, V. (2021). GAD-7, GAD-2, and GAD-mini: Psychometric properties and norms of university students in the United States. General Hospital Psychiatry, 69, 61–66. 10.1016/j.genhosppsych.2021.01.002

Carpita, B., Nardi, B., Muti, D., Battaglini, S., Parri, F., Giovannoni, F., Cerofolini, G., Bonelli, C., Cremone, I. M., Massimetti, G., Di Vincenzo, M., Pini, S., Pellecchia, E., Fiorillo, A., & Dell’Osso, L. (2026). Hikikomori-like social withdrawal mediates the relationship between autistic traits and pathological video game use among university students. International Journal of Social Psychiatry, 1388–1399. 10.1177/00207640251405460

Cekic, S., Bediou, B., Achab, S., Rich, M., Green, C. S., & Bavelier, D. (2024). Going beyond video game consumption when considering Internet Gaming Disorder. Comprehensive Psychiatry, 133, Article 152500. 10.1016/j.comppsych.2024.152500

Cheung, T. C. K., Reza, T., B., Pereira, C. F., Mukhi, S., & Niemeier, M. (2023). Limited reliability and validity of the Autism Spectrum Quotient short form (AQ-10) to screen autistic traits in undergraduate students. Journal of Autism and Developmental Disorders, 53(7), 2919–2920. 10.1007/s10803-022-05889-1

Coelho, M. D. M. F., Cavalcante, V. M. V., Castro, T. H., Oriá, M. O. B., Beserra, E. P., Gubert, F. D. A., Martins, M. C., Marques, M. B., Nogueira, P. S. F., Coutinho, J. F. V., & Barbosa, R. G. B. (2025). Confirmatory factor analysis of the internet addiction test in university students. Revista Cuidarte, 16(2), Article e4593. 10.15649/cuidarte.4593

Concerto, C., Rodolico, A., Avanzato, C., Fusar-Poli, L., Signorelli, M. S., Battaglia, F., & Aguglia, E. (2021). Autistic traits and Attention-Deficit Hyperactivity Disorder symptoms predict the severity of internet gaming disorder in an Italian adult population. Brain Sciences, 11(6), Article 774. 10.3390/brainsci11060774

de-Sola, J., Talledo, H., Rodríguez De Fonseca, F., & Rubio, G. (2017). Prevalence of problematic cell phone use in an adult population in Spain as assessed by the Mobile Phone Problem Use Scale (MPPUS). PLOS ONE, 12(8), Article e0181184. 10.1371/journal.pone.0181184

Dhira, T. A., Rahman, M. A., Sarker, A. R., & Mehareen, J. (2021). Validity and reliability of the Generalized Anxiety Disorder-7 (GAD-7) among university students of Bangladesh. PLOS ONE, 16(12), Article e0261590. 10.1371/journal.pone.0261590

Elhai, J. D., Casale, S., & Montag, C. (2025). Worry and fear of missing out are associated with problematic smartphone and social media use severity. Journal of Affective Disorders, 379, 258–265. 10.1016/j.jad.2025.03.062

Enders, C., & Bandalos, D. (2001). The relative performance of full information maximum likelihood estimation for missing data in structural equation models. Structural Equation Modeling: A Multidisciplinary Journal, 8(3), 430–457. 10.1207/S15328007SEM0803_5

Fatséas, M., Hurmic, H., Serre, F., Debrabant, R., Daulouède, J.-P., Denis, C., & Auriacombe, M. (2016). Addiction severity pattern associated with adult and childhood Attention Deficit Hyperactivity Disorder (ADHD) in patients with addictions. Psychiatry Research, 246, 656–662. 10.1016/j.psychres.2016.10.071

Fegert, J. M., Gottschalk, G., Schneider, R., Sitarski, E., Sounderajah, V., & Graham, G. (2025). Navigating life transitions and mental wellbeing in the digital age: A call for stakeholders to embrace innovation and collaboration. Child and Adolescent Psychiatry and Mental Health, 19(1), Article 67. 10.1186/s13034-025-00932-2

Fraiwan, M., & Almomani, F. (2025). Internet Gaming Disorder: Prevalence and associated factors among university students. Acta Psychologica, 260, Article 105690. 10.1016/j.actpsy.2025.105690

Garcha, J., & Smith, A. P. (2023). Associations between autistic and ADHD traits and the well-being and mental health of university students. Healthcare, 12(1), Article 14. 10.3390/healthcare12010014

González-Bueso, V., Santamaría, J., Fernández, D., Merino, L., Montero, E., & Ribas, J. (2018). Association between Internet Gaming Disorder or pathological video-game use and comorbid psychopathology: A comprehensive review. International Journal of Environmental Research and Public Health, 15(4), Article 668. 10.3390/ijerph15040668

Grant, J. E., & Chamberlain, S. R. (2021). Autistic traits in young adults who gamble. CNS Spectrums, 26(6), 637–642. 10.1017/S1092852920001571

Hegarty, D., Buchanan, B., Baker, S., Bartholomew, E., & Smyth, C. (2024). Adult ADHD self-report scale (ASRS): Updated scoring and norms. https://novopsych.com/wp-content/uploads/2024/11/Adult-ADHD-Self-Report-Scale-ASRS-Updated-Scoring-and-Norms.pdf

Ho, R. C., Zhang, M. W., Tsang, T. Y., Toh, A. H., Pan, F., Lu, Y., Cheng, C., Yip, P. S., Lam, L. T., Lai, C.-M., Watanabe, H., & Mak, K.-K. (2014). The association between internet addiction and psychiatric co-morbidity: A meta-analysis. BMC Psychiatry, 14(1), Article 183. 10.1186/1471-244X-14-183

Kalınkara, Y., & Talan, T. (2025). Psychological balances in the digital world: Dynamic relationships among social media addiction, depression, anxiety, academic self-efficacy, general belongingness, and life satisfaction. The Journal of Genetic Psychology, 186(2), 85–113. 10.1080/00221325.2024.2400342

Kaya, F., Bayzan, Ş., Bhais, A. Z. B., & Gün, A. (2025). Lonely, anxious, and unhappy: The psychosocial toll of digital game addiction in university students. International Journal of Mental Health and Addiction, 24(4), 3510–3535. 10.1007/s11469-025-01568-1

Kessler, R. C., Adler, L., Ames, M., Demler, O., Faraone, S., Hiripi, E., Howes, M. J., Jin, R., Secnik, K., Spencer, T., Ustun, T. B., & Walters, E. E. (2005). The World Health Organization adult ADHD self-report scale (ASRS): A short screening scale for use in the general population. Psychological Medicine, 24(2), 245–256. 10.1017/S0033291704002892

Kim, S., Champion, K. E., Gardner, L. A., Teesson, M., Newton, N. C., & Gainsbury, S. M. (2022). The directionality of anxiety and gaming disorder: An exploratory analysis of longitudinal data from an Australian youth population. Frontiers in Psychiatry, 13, Article 1043490. 10.3389/fpsyt.2022.1043490

Kofler, M. J., Soto, E. F., Singh, L. J., Harmon, S. L., Jaisle, E. M., Smith, J. N., Feeney, K. E., & Musser, E. D. (2024). Executive function deficits in attention-deficit/hyperactivity disorder and autism spectrum disorder. Nature Reviews Psychology, 3(10), 701–719. 10.1038/s44159-024-00350-9

Köse, S., & Umut, İ. (2024). The relationship between autistic traits, internet addiction, perceived social support and life satisfaction. Archives of Neuropsychiatry. 10.29399/npa.28752

Lewczuk, K., Marcowski, P., Wizła, M., Gola, M., Nagy, L., Koós, M., Kraus, S. W., Demetrovics, Z., Potenza, M. N., Ballester-Arnal, R., Batthyány, D., Bergeron, S., Billieux, J., Briken, P., Burkauskas, J., Cárdenas-López, G., Carvalho, J., Castro-Calvo, J., Chen, L., … Bőthe, B. (2024). Cross-cultural adult ADHD assessment in 42 countries using the Adult ADHD Self-Report Scale Screener. Journal of Attention Disorders, 28(4), 512–530. 10.1177/10870547231215518

Liu, X., Gui, Z., Chen, Z.-M., Feng, Y., Wu, X., Su, Z., Cheung, T., Ungvari, G. S., Liu, X.-C., Yan, Y.-R., Ng, C. H., & Xiang, Y.-T. (2025). Global prevalence of internet addiction among university students: A systematic review and meta-analysis. Current Opinion in Psychiatry, 38(3), 182–199. 10.1097/YCO.0000000000000994

Lyvers, M., Dark, S., Jaguru, I., & Thorberg, F. A. (2024). Adult symptoms of ASD and ADHD in relation to alcohol use: Potential roles of transdiagnostic features. Alcohol, 120, 109–117. 10.1016/j.alcohol.2024.03.011

Moon, S. J., Hwang, J. S., Kim, J. Y., Shin, A. L., Bae, S. M., & Kim, J. W. (2018). Psychometric properties of the Internet Addiction Test: A systematic review and meta-analysis. Cyberpsychology, Behavior, and Social Networking, 21(8), 473–484. 10.1089/cyber.2018.0154

Nankoo, M. M. A., Palermo, R., Bell, J. A., & Pestell, C. M. (2019). Examining the rate of self-reported ADHD-related traits and endorsement of depression, anxiety, stress, and autistic-like traits in Australian university students. Journal of Attention Disorders, 23(8), 869–886. 10.1177/1087054718758901

Oviedo-Trespalacios, O., Nandavar, S., Newton, J. D. A., Demant, D., & Phillips, J. G. (2019). Problematic use of mobile phones in Australia…Is it getting worse? Frontiers in Psychiatry, 10, Article 105. 10.3389/fpsyt.2019.00105

R Core Team. (2025). R: A language and environment for statistical computing (Version 4.5.2) [Computer software]. R Foundation for Statistical Computing. https://www.R-project.org/

Rosseel, Y. (2012). lavaan: An R package for structural equation modeling: 1–36. Journal of Statistical Software, 48(2). 10.18637/jss.v048.i02

Rosseel, Y., Jorgensen, T. D., & De Wilde, L. (2025). *lavaan: Latent variable analysis* (Version 0.6-20) [Computer software]. 10.32614/CRAN.package.lavaan

Salari, N., Zarei, H., Hosseinian-Far, A., Rasoulpoor, S., Shohaimi, S., & Mohammadi, M. (2025). The global prevalence of social media addiction among university students: A systematic review and meta-analysis. Journal of Public Health, 33(1), 223–236. 10.1007/s10389-023-02012-1

Salman, I., Bajwa, R. S., & Yunus, A. (2025). Association between digital media use and biopsychosocial health of students. Journal of Technology in Behavioral Science, 11, 463–471. 10.1007/s41347-025-00531-0

Sayili, U., Kara, B., Toplu, F. S., Yilmaz Kose, S. N., Sak, K., Kilickan, F. D. B., & Can, G. (2025). Do scales measuring internet addiction give compatible results? Comparison of two internet addiction scales among medical students. BMC Public Health, 25(1), Article 3560. 10.1186/s12889-025-24695-9

Senkowski, D., Ziegler, T., Singh, M., Heinz, A., He, J., Silk, T., & Lorenz, R. C. (2024). Assessing inhibitory control deficits in adult ADHD: A systematic review and meta-analysis of the stop-signal task. Neuropsychology Review, 34(2), 548–567. 10.1007/s11065-023-09592-5

Shiferaw, B. D., Tang, J., Wang, Y., Wang, Y., Wang, Y., Mackay, L. E., Luo, Y., Yan, N., Shen, X., Zhou, T., Zhu, Y., Cai, J., Wang, Q., Yan, W., Gao, X., Pan, H., & Wang, W. (2025). Impact of digital addiction on youth health: A systematic review and meta-analysis. Journal of Behavioral Addictions, 14(3), 1129–1158. 10.1556/2006.2025.00081

Singh, A. K., & Singh, P. K. (2019). Digital addiction: A conceptual overview. Library Philosophy and Practice. https://scholarworks.sjsu.edu/libphilprac/3538/

Slack, J. D., Mctigue, B., Mackenzie, H., Flack, M., & Caudwell, K. M. (2025). A mediation analysis of Autistic-like traits and gaming motivations on problem gaming symptoms: Are the effects of social and escape motives the same? Addictive Behaviors Reports, 22, Article 100625. 10.1016/j.abrep.2025.100625

Southon, C. (2022). The relationship between executive function, neurodevelopmental disorder traits, and academic achievement in university students. Frontiers in Psychology, 13, Article 958013. 10.3389/fpsyg.2022.958013

Spitzer, R. L., Kroenke, K., Williams, J. B. W., & Löwe, B. (2006). A brief measure for assessing generalized anxiety disorder: The GAD-7. Archives of Internal Medicine, 166(10), 1092–1097. 10.1001/archinte.166.10.1092

Stocker, R., Tran, T., Hammarberg, K., Nguyen, H., Rowe, H., & Fisher, J. (2021). Patient Health Questionnaire 9 (PHQ-9) and General Anxiety Disorder 7 (GAD-7) data contributed by 13,829 respondents to a national survey about COVID-19 restrictions in Australia. Psychiatry Research, 298, Article 113792. 10.1016/j.psychres.2021.113792

Sulla, F., Camia, M., Scorza, M., Giovagnoli, S., Padovani, R., & Benassi, E. (2023). The moderator effect of subthreshold autistic traits on the relationship between quality of life and internet addiction. Healthcare, 11(2), Article 186. 10.3390/healthcare11020186

Townes, P., Liu, C., Panesar, P., Devoe, D., Lee, S. Y., Taylor, G., Arnold, P. D., Crosbie, J., & Schachar, R. (2023). Do ASD and ADHD have distinct executive function deficits? A systematic review and meta-analysis of direct comparison studies. Journal of Attention Disorders, 27(14), 1571–1582. 10.1177/10870547231190494

Tülübaş, T., Karakose, T., & Papadakis, S. (2023). A holistic investigation of the relationship between digital addiction and academic achievement among students. European Journal of Investigation in Health, Psychology and Education, 13(10), 2006–2034. 10.3390/ejihpe13100143

Van Rooij, A. J., Schoenmakers, T. M., & Van De Mheen, D. (2017). Clinical validation of the C-VAT 2.0 assessment tool for gaming disorder: A sensitivity analysis of the proposed DSM-5 criteria and the clinical characteristics of young patients with ‘video game addiction.’ Addictive Behaviors, 64, 269–274. 10.1016/j.addbeh.2015.10.018

Van Rooij, A. J., Schoenmakers, T. M., Van Den Eijnden, R. J. J. M., Vermulst, A. A., & Van De Mheen, D. (2012). Video Game Addiction Test: Validity and psychometric characteristics. Cyberpsychology, Behavior, and Social Networking, 15(9), 507–511. 10.1089/cyber.2012.0007

von Rhein, D., Cools, R., Zwiers, M. P., Van Der Schaaf, M., Franke, B., Luman, M., Oosterlaan, J., Heslenfeld, D. J., Hoekstra, P. J., Hartman, C. A., Faraone, S. V., van Rooij, D., van Dongen, E. V., Lojowska, M., Mennes, M., & Buitelaar, J. (2015). Increased neural responses to reward in adolescents and young adults with attention-deficit/hyperactivity disorder and their unaffected siblings. Journal of the American Academy of Child & Adolescent Psychiatry, 54(5), 394–402. 10.1016/j.jaac.2015.02.012

Waite, R., Buchanan, T., & Leahy, M. (2022). Assessing ADHD symptoms among young adults in the university with the ASRS v1.1: Examining associations with social anxiety and self-efficacy. International Journal of Disability, Development and Education, 69(5), 1631–1644. 10.1080/1034912X.2020.1825643

Wang, Y., & Abdullah, M. N. L. Y. (2026). A meta-analytic review of smartphone addiction prevalence in Chinese college students. Current Psychology, 45(5), Article 548. 10.1007/s12144-026-09160-z

Wei, Z., Hassan, N. C., Hassan, S. A., Ismail, N., Gu, X., & Dong, J. (2025). Psychometric validation of Young’s Internet Addiction Test among Chinese undergraduate students. PLOS One, 20(4), Article e0320641. 10.1371/journal.pone.0320641

Wilens, T. E., Martelon, M., Joshi, G., Bateman, C., Fried, R., Petty, C., & Biederman, J. (2011). Does ADHD predict substance-use disorders? A 10-year follow-up study of young adults with ADHD. Journal of the American Academy of Child & Adolescent Psychiatry, 50(6), 543–553. 10.1016/j.jaac.2011.01.021

Yankouskaya, A., Liebherr, M., & Ali, R. (2025). Can ChatGPT be addictive? A call to examine the shift from support to dependence in AI conversational large language models. Human-Centric Intelligent Systems, 5(1), 77–89. 10.1007/s44230-025-00090-w

Young, K. S. (1998). Caught in the net: How to recognize the signs of Internet addiction--and a winning strategy for recovery. J. Wiley.

Zhou, S., Chen, Z., & Liu, Y. (2024). The relationship between autistic traits and problematic smartphone use in adolescents: The serial mediating role of anxiety and executive dysfunction. BMC Psychology, 12(1), Article 683. 10.1186/s40359-024-02180-z

