## Supplementary for "Generalised anxiety as a mediator linking ADHD symptoms and autistic traits to digital addictions in Australian emerging adults"

#### Author Note

Matthew J. Giblett https://orcid.org/0009-0006-0400-6575

Brooke Andrew https://orcid.org/0000-0001-5532-799X

Penelope A. Lind https://orcid.org/0000-0002-3887-2598

Cara Jaklich is now at the Department of Special Education, The University of Texas at Austin.

We have no known conflicts of interests to disclose.

Correspondence concerning this article should be addressed to Matthew J. Giblett, Psychiatric Genetics, Brain and Mental Health Research Program, QIMR Berghofer, 300 Herston Road, Herston, Brisbane, Queensland 4006, Australia.

### Results

#### Missing data analysis

Analyses were performed using R (version 4.5.2, R Core Team, 2025) with the *lavaan* package (version 0.6-21, Rosseel, 2012; Rosseel et al., 2025) for confirmatory factor analysis (CFA) and path analysis. We examined all participants ($n$ = 222) and variables for missing data prior to conducting main analyses. At the item-level, a total of 3,153 out of 18,648 (16.9%) data points were missing. When computing the total scores for each measure, we used prorating only when participants were missing responses to no more than 20% of items (e.g., fewer than three items on the Autism Quotient–10 [AQ-10], Allison et al., 2012). When missingness exceeded this threshold, total scores were not computed and were coded as missing. As a result, we retained 11 scale-level datapoints that would otherwise have been coded as missing. After computing total scores, missing data were present for all measures except sex, with 63 participants (28.4%) missing at least one. At the scale-level, a total of 225 out of 1,332 (16.9%) data points were missing. Scale-level missingness ranged from 10.4% to 23.4%, with Internet Addiction Test (IAT, Young, 1998) having the least and Video Game Addiction Test (VAT, Van Rooij et al., 2012) the most.

We performed a Little's Missing Completely at Random (MCAR) test (Little, 1988) with the *naniar* package (Tierney & Cook, 2023) to assess whether missing data were MCAR. The test included all primary outcome and sociodemographic variables hypothesised to be related to missingness. The test was significant, $\chi^2(101) = 127.45$, $p = .039$, indicating that the assumption of MCAR was not supported. This result suggests that missingness may be Missing at Random (MAR) or Missing Not at Random (MNAR).

To investigate any potential mechanisms of missingness, we performed additional follow-up analyses comparing participants with no missing data (complete cases) to those with missing data (incomplete cases). Table S1 compares categorical variables between the two groups using Pearson's Chi-squared test, and Table S2 compares continuous variables using Welch's Two Sample *t*-test. Results revealed that non–Queensland University of Technology (QUT) School of Psychology students and participants with a university degree of any level had significantly higher proportions of missing data than QUT School of Psychology students and those without a university degree, respectively. Incomplete participants were also significantly older than complete participants. These findings indicate systematic patterns of missingness related to observed variables, which is consistent with MAR (Bhaskaran & Smeeth, 2014). Consequently, missing data was handled using Full Information Maximum Likelihood (FIML, Enders & Bandalos, 2001) implemented in R using the *lavaan* package, allowing all available data to contribute to parameter estimation under the assumption of MAR.

**Table S1**

*Comparison of Demographic Characteristics between Participants with Complete and Incomplete Data.*

| Characteristic | Missingness Level | | $\chi^2$ | *p* |
|---|---|---|---|---|
| | Incomplete ($n$ = 63) | Complete ($n$ = 159) | | |
| Sex, *n* (%) | | | 0.80 | .473 |
| Female | 46 (73) | 125 (79) | | |
| Male | 17 (27) | 34 (21) | | |
| QUT School of Psychology student at survey, *n* (%) | | | 16.14 | <.001*** |
| No | 24 (39) | 22 (14) | | |
| Yes | 38 (61) | 134 (86) | | |
| Highest level of education, *n* (%) | | | 6.64 | .036* |

| Characteristic | Missingness Level | | $\chi^2$ | *p* |
|---|---|---|---|---|
| | Incomplete (*n* = 63) | Complete (*n* = 159) | | |
| Certificate or Diploma | 9 (14) | 23 (14) | | |
| Senior High School | 40 (63) | 121 (76) | | |
| University degree (any level) | 14 (22) | 15 (9.4) | | |
| Employment, *n* (%) | | | 0.13 | .832 |
| Employed | 30 (48) | 71 (45) | | |
| Not employed | 33 (52) | 87 (55) | | |
| Income, *n* (%) | | | 1.47 | .480 |
| ≤ $39,999 | 11 (17) | 22 (14) | | |
| ≥ $40,000 | 16 (25) | 53 (33) | | |
| Income unknown / not disclosed | 36 (57) | 84 (53) | | |

*Note.* Incomplete = participants missing any data; Complete = participants missing no data; QUT = Queensland University of Technology.

* $p < .05$. ** $p < .01$. *** $p < .001$.

**Table S2**

*Comparison of Continuous Variables Between Complete and Incomplete Participants*

| Characteristic | Missingness Level | | *t* | *p* |
|---|---|---|---|---|
| | Incomplete (*n* = 63) | Complete (*n* = 159) | | |
| Age at survey, years, *M* (*SD*) | 20.9 (2.8) | 19.6 (2.9) | 3.17 | .002** |
| AQ-10, *M* (*SD*) | 21.2 (4.3) | 20.9 (3.4) | 0.25 | .804 |
| ASRS-S, *M* (*SD*) | 9.7 (4.8) | 9.3 (4.2) | 0.38 | .707 |
| GAD-7, *M* (*SD*) | 6.2 (5.6) | 5.8 (5.1) | 0.32 | .752 |
| IAT, *M* (*SD*) | 17.6 (12.1) | 17.2 (10.1) | 0.19 | .851 |

| Characteristic | Missingness Level | | *t* | *p* |
|---|---|---|---|---|
| | Incomplete (*n* = 63) | Complete (*n* = 159) | | |
| MPPUS, *M* (*SD*) | 100.0 (40.1) | 103.9 (42.2) | −0.42 | .680 |
| VAT, *M* (*SD*) | 18.0 (17.3) | 8.6 (10.0) | 1.79 | .103 |

*Note*. Complete = participants missing no data; Incomplete = participants missing any data; AQ-10 = Autism Spectrum Quotient-10; ASRS-S = Adult ADHD Self-Report Scale v1.1 screener (Kessler et al., 2005); GAD-7 = Generalised Anxiety Disorder-7 (Spitzer et al., 2006); IAT = Internet Addiction Test; MPPUS = Mobile Phone Problem Use Scale; VAT = Video Game Addiction Test.

* $p < .05$. ** $p < .01$. *** $p < .001$.

#### Assumption checks

##### *Normality*

We assessed each continuous variable for univariate normality using the Anderson–Darling test implemented in the *nortest* package (Gross & Ligges, 2006). Table S3 shows the skewness, kurtosis, and normality test results for each variable, and Figure S1 shows histograms for each continuous variable. These results reveal that all variables are not normally distributed. Multivariate normality was assessed using the Henze-Zirkler test (HZ, Henze & Zirkler, 1990), implemented via the *MVN* package (Korkmaz et al., 2014). The test indicated a significant departure from multivariate normality, $HZ = 1.6$, $p < .001$. See Figure S2 for a Q-Q plot. Given the departures from univariate and multivariate normality, nonparametric bootstrapping (k = 5,000 samples) was used to estimate confidence intervals for indirect effects, as it makes no distributional assumptions and is robust to non-normality (Hayes, 2022).

**Table S3**

*Univariate Normality Diagnostics for Study Variables*

| Variable | *N* | Skew | Kurtosis | AD | *p* |
|---|---|---|---|---|---|
| AQ-10 | 186 | 0.51 | 0.76 | 1.13 | .006** |
| ASRS-S | 186 | 0.20 | 0.46 | 0.82 | .033* |
| GAD-7 | 186 | 1.31 | 1.38 | 7.22 | < .001*** |
| IAT | 199 | 0.89 | 0.65 | 3.51 | < .001*** |
| MPPUS | 180 | 0.22 | -0.41 | 0.74 | .054 |
| VAT | 170 | 1.40 | 1.84 | 9.38 | < .001*** |

*Note.* AD = Anderson-Darling Normality Test; AQ-10 = Autism Spectrum Quotient-10; ASRS-S = Adult ADHD Self-Report Scale v1.1 screener; GAD-7 = Generalised Anxiety Disorder-7; IAT = Internet Addiction Test; MPPUS = Mobile Phone Problem Use Scale; VAT = Video Game Addiction Test.

* $p < .05$. ** $p < .01$. *** $p < .001$.

**Figure S1**

*Histograms of (a) Autism Spectrum Quotient (AQ-10), (b) Adult ADHD Self-Report Scale v1.1 screener (ASRS-S), (c) Generalised Anxiety Disorder-7 (GAD-7), (d) Internet Addiction Test (IAT), (e) Mobile Phone Problem Use Scale (MPPUS), and (f) Video Game Addiction Test (VAT) Total Scores*

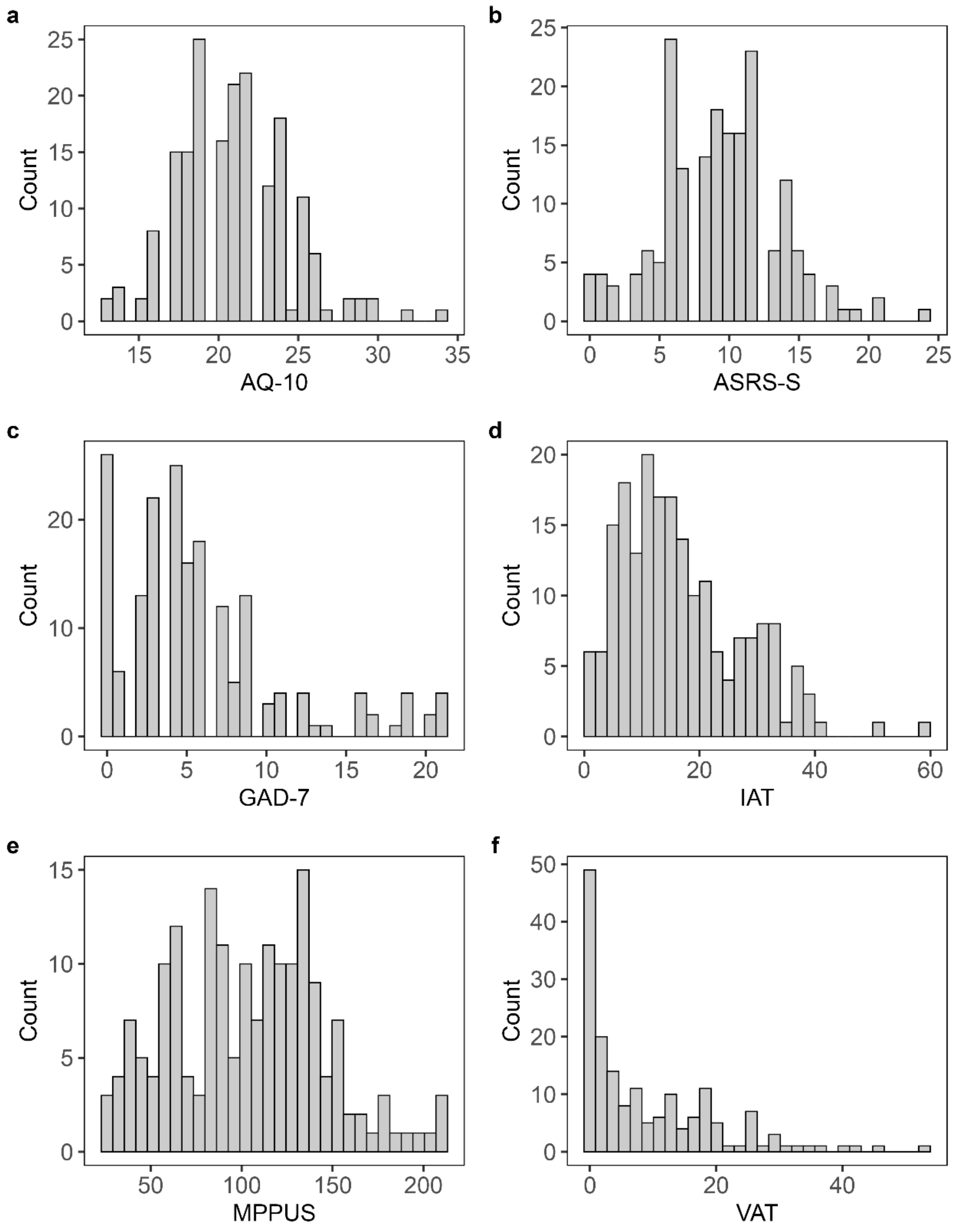

**Figure S2**

*Q-Q Plot of Squared Mahalanobis Distance against Chi-Square Quantiles*

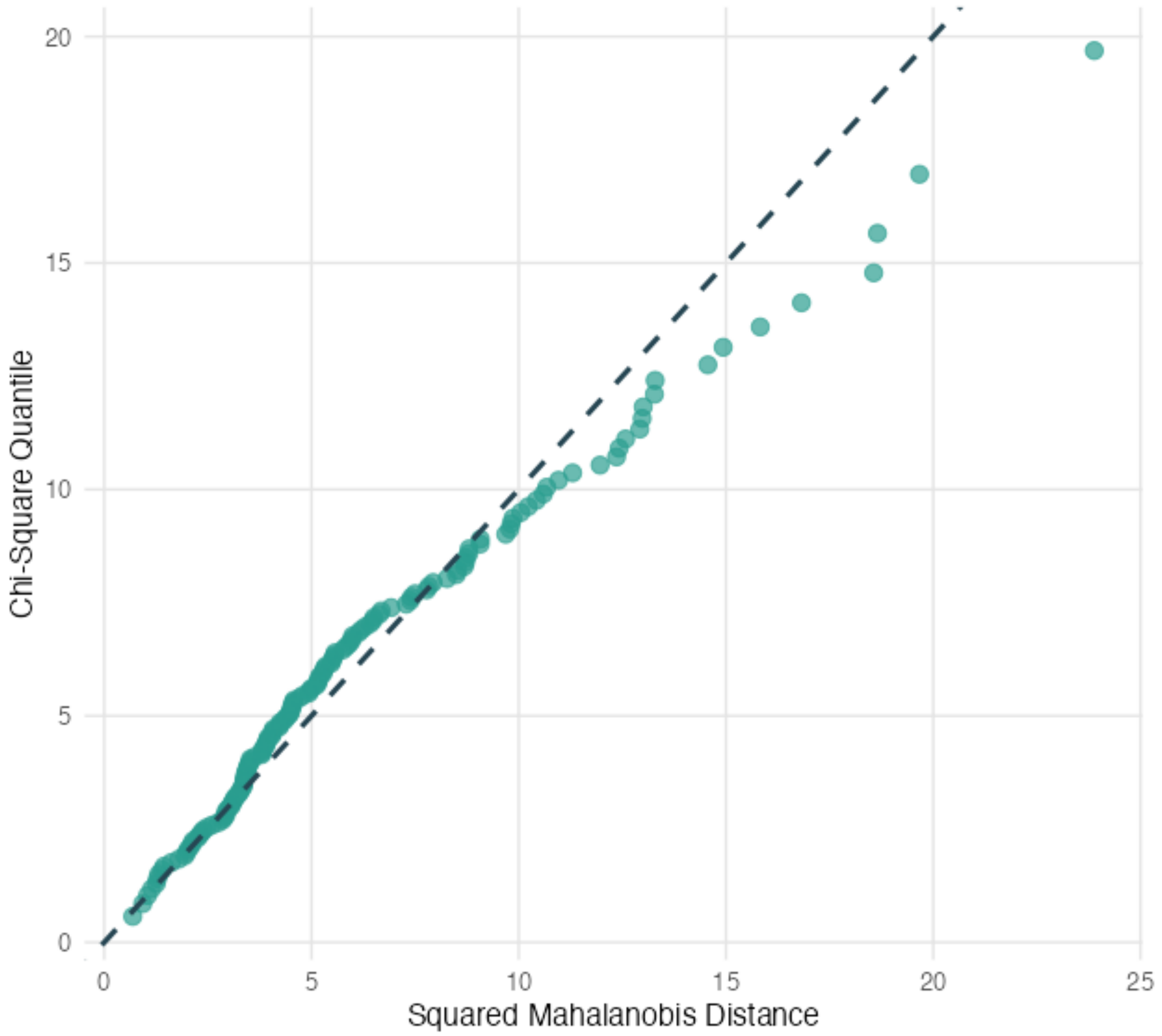

***Multicollinearity***

Multicollinearity was assessed for each predictor of each outcome variable using variance inflation factors (VIF), implemented via the *car* package (Fox & Weisberg, 2019). Table S4 presents the results of these analyses and reveals no problematic multicollinearity.

**Table S4**

*Variance Inflation Factors for each Predictor of GAD-7, IAT, MPPUS, and VAT*

| Predictor | GAD-7 [a] | IAT | MPPUS | VAT |
|---|---|---|---|---|
| AQ-10 | 1.04 | 1.06 | 1.07 | 1.09 |
| ASRS-S | 1.04 | 1.31 | 1.30 | 1.31 |
| Sex [b] | 1.00 | 1.06 | 1.07 | 1.11 |
| GAD-7 [a] | — | 2.24 | 2.32 | 2.40 |
| Sex × GAD-7 | — | 1.82 | 1.92 | 1.98 |

*Note.* AQ-10 = Autism Spectrum Quotient-10; ASRS-S = Adult ADHD Self-Report Scale v1.1 screener; GAD-7 = Generalised Anxiety Disorder-7; IAT = Internet Addiction Test; MPPUS = Mobile Phone Problem Use Scale; VAT = Video Game Addiction Test.

[a] GAD-7 was mean-centred across all analyses to reduce multicollinearity with the interaction term.

[b] Sex was coded –1 = male, 1 = female.

***Linearity***

Linearity among the predictors and the outcomes was assessed using component-plus-residual plots (CR plot) whilst controlling for all other predictors in the model implemented via the *car* package. As some models included the Sex × GAD-7 interaction term, CR plots for each of the digital addictions were run at each level of sex as well as in the full sample without the interaction (Nahhas, 2024). These plots indicated minor deviations in linearity.

##### *Outliers*

Outliers were assessed using robust Mahalanobis distance, generalised Cook's distance, and goodness of fit distance, implemented in the *faoutlier* package (Chalmers & Flora, 2015). There were no consistent outliers across models and methods, so all outliers were retained to avoid excluding genuine variation in the data.

#### Path Analyses with Sex × GAD-7 Interaction

See Table S5 for the fit indices of each of the three interaction models and Table S6 for the path estimates of each predictor of IAT, MPPUS, and VAT in their respective models, including the total and indirect effects of AQ-10 and ASRS-S through GAD-7. The proportion of variance explained ($R^2$) for GAD-7 was 0.26, 0.25 for IAT, 0.22 for MPPUS, and 0.40 for VAT.

**Table S5**

*Fit Indices of Path Analysis Models Examining Associations among AQ-10, ASRS-S, Participant Sex, GAD-7, Sex × GAD-7, and Digital Addiction Outcomes*

| Model | Chi-Square Goodness of Fit | | | Incremental Fit | | Absolute Fit | | |
|---|---|---|---|---|---|---|---|---|
| | $\chi^2$ | *df* | *p* | CFI | TLI | RMSEA | 95% CI | SRMR |
| IAT | 109.68 | 5 | < .001*** | 0.45 | -0.53 | 0.31 | [0.26, 0.36] | 0.15 |
| MPPUS | 110.65 | 5 | < .001*** | 0.47 | -0.48 | 0.31 | [0.26, 0.36] | 0.14 |
| VAT | 109.94 | 5 | < .001*** | 0.53 | -0.31 | 0.31 | [0.26, 0.36] | 0.16 |

*Note*. CI = confidence interval; CFI = Comparative Fit Index; TLI = Tucker-Lewis Index; RMSEA = Root Mean Square Error of Approximation; SRMR = Standardized Root Mean Square Residual; IAT = Internet Addiction Test; MPPUS = Mobile Phone Problem Use Scale; VAT = Video Game Addiction Test.

**Table S6**

*Parameter Estimates of Path Analyses Examining Associations among AQ-10, ASRS-S, Participant Sex, GAD-7, Sex × GAD-7, and Digital Addictions*

| Parameter | IAT | | | | | MPPUS | | | | | VAT | | | | |
|---|---|---|---|---|---|---|---|---|---|---|---|---|---|---|---|
| | *B* | 95% CI [a] | *SE* [a] | β | *p* FDR [b] | *B* | 95% CI [a] | *SE* [a] | β | *p* FDR [b] | *B* | 95% CI [a] | *SE* [a] | β | *p* FDR [b] |
| **Covariances** | | | | | | | | | | | | | | | |
| AQ-10 ↔ ASRS-S | 2.96 | [0.4, 5.74] | 1.37 | 0.20 | .091 | 2.88 | [0.35, 5.65] | 1.35 | 0.19 | .091 | 2.95 | [0.4, 5.73] | 1.36 | 0.20 | .091 |
| Sex ↔ Sex × GAD-7 [c] | −0.28 | [−0.86, 0.32] | 0.30 | −0.07 | .376 | −0.30 | [−0.87, 0.3] | 0.30 | −0.07 | .376 | −0.28 | [−0.85, 0.33] | 0.30 | −0.07 | .376 |
| **Direct effects on GAD-7** | | | | | | | | | | | | | | | |
| AQ-10 | 0.20 | [0.04, 0.36] | 0.08 | 0.14 | .017* | 0.19 | [0.04, 0.36] | 0.08 | 0.13 | .017* | 0.20 | [0.04, 0.36] | 0.08 | 0.14 | .017* |
| ASRS-S | 0.54 | [0.38, 0.7] | 0.08 | 0.45 | < .001*** | 0.54 | [0.38, 0.7] | 0.08 | 0.44 | < .001*** | 0.55 | [0.38, 0.7] | 0.08 | 0.45 | < .001*** |
| Sex [c] | 0.71 | [0.01, 1.41] | 0.36 | 0.12 | .102 | 0.75 | [0.05, 1.45] | 0.36 | 0.12 | .091 | 0.68 | [−0.03, 1.38] | 0.36 | 0.11 | .108 |
| **Direct effects on digital addiction** | | | | | | | | | | | | | | | |
| GAD-7 | 0.57 | [0.03, 1.14] | 0.29 | 0.27 | .069 | 1.31 | [−0.54, 2.99] | 0.90 | 0.16 | .145 | 0.82 | [0.28, 1.33] | 0.27 | 0.38 | .007** |
| AQ-10 | 0.47 | [0.05, 0.91] | 0.22 | 0.15 | .089 | 0.56 | [−1.04, 2.28] | 0.84 | 0.05 | .503 | 0.38 | [−0.08, 0.8] | 0.22 | 0.12 | .141 |
| ASRS-S | 0.52 | [0.06, 0.95] | 0.23 | 0.20 | .027* | 2.61 | [1.12, 4.06] | 0.74 | 0.26 | .001** | 0.39 | [0.04, 0.74] | 0.18 | 0.15 | .027* |
| Sex [c] | −1.50 | [−3.77, 0.66] | 1.12 | −0.12 | .275 | 12.90 | [5.53, 19.93] | 3.68 | 0.26 | .003** | −5.51 | [−7.64, −3.28] | 1.11 | −0.41 | < .001*** |
| Sex × GAD-7 [c] | −0.36 | [−0.9, 0.16] | 0.27 | −0.17 | .275 | −0.26 | [−1.77, 1.38] | 0.81 | −0.03 | .752 | −0.25 | [−0.75, 0.27] | 0.26 | −0.11 | .376 |

| Parameter | IAT | | | | | MPPUS | | | | | VAT | | | | |
|---|---|---|---|---|---|---|---|---|---|---|---|---|---|---|---|
| | *B* | 95% CI [a] | *SE* [a] | β | *p* FDR [b] | *B* | 95% CI [a] | *SE* [a] | β | *p* FDR [b] | *B* | 95% CI [a] | *SE* [a] | β | *p* FDR [b] |
| **Indirect effects on digital addiction through GAD-7** | | | | | | | | | | | | | | | |
| AQ-10 | 0.11 | [0, 0.3] | 0.08 | 0.04 | .223 | 0.25 | [−0.1, 0.71] | 0.21 | 0.02 | .223 | 0.16 | [0.02, 0.36] | 0.09 | 0.05 | .193 |
| ASRS-S | 0.31 | [0.01, 0.64] | 0.16 | 0.12 | .079 | 0.70 | [−0.28, 1.77] | 0.51 | 0.07 | .166 | 0.45 | [0.14, 0.78] | 0.17 | 0.17 | .021* |
| **Total effects on digital addiction** | | | | | | | | | | | | | | | |
| AQ-10 | 0.59 | [0.14, 1.05] | 0.23 | 0.19 | .030* | 0.82 | [−0.82, 2.51] | 0.83 | 0.07 | .326 | 0.54 | [0.06, 0.99] | 0.23 | 0.17 | .033* |
| ASRS-S | 0.83 | [0.39, 1.27] | 0.22 | 0.32 | < .001*** | 3.32 | [1.79, 4.73] | 0.75 | 0.33 | < .001*** | 0.84 | [0.39, 1.27] | 0.22 | 0.32 | < .001*** |

*Note*. CI = confidence interval; AQ-10 = Autism Spectrum Quotient-10; ASRS-S = Adult ADHD Self-Report Scale v1.1 screener; GAD-7 = Generalised Anxiety Disorder-7; IAT = Internet Addiction Test; MPPUS = Mobile Phone Problem Use Scale; VAT = Video Game Addiction Test.

[a] *SE*s and CIs were obtained via nonparametric bootstrapping (5,000 resamples).

[b] *p* FDR values are Benjamini–Hochberg false discovery rate (FDR) adjusted *p*-values.

[c] Sex was coded –1 = male, 1 = female.

* $p < .05$. ** $p < .01$. *** $p < .001$

**References**

Allison, C., Auyeung, B., & Baron-Cohen, S. (2012). Toward brief “red flags” for autism screening: The short Autism Spectrum Quotient and the short quantitative checklist in 1,000 cases and 3,000 controls. *Journal of the American Academy of Child & Adolescent Psychiatry*, *51*(2), 202–212. https://doi.org/10.1016/j.jaac.2011.11.003

Bhaskaran, K., & Smeeth, L. (2014). What is the difference between missing completely at random and missing at random? *International Journal of Epidemiology*, *43*(4), 1336–1339. https://doi.org/10.1093/ije/dyu080

Chalmers, R. P., & Flora, D. B. (2015). faoutlier: An R package for detecting influential cases in exploratory and confirmatory factor analysis. *Applied Psychological Measurement*, *39*(7), 573–574. https://doi.org/10.1177/0146621615597894

Enders, C., & Bandalos, D. (2001). The relative performance of full information maximum likelihood estimation for missing data in structural equation models. *Structural Equation Modeling: A Multidisciplinary Journal*, *8*(3), 430–457. https://doi.org/10.1207/S15328007SEM0803_5

Fox, J., & Weisberg, S. (2019). *An R companion to applied regression* (3rd ed.). Sage. https://www.john-fox.ca/Companion/

Gross, J., & Ligges, U. (2006). *nortest: Tests for Normality* (Version 1.0-4) [Computer software]. https://doi.org/10.32614/cran.package.nortest

Hayes, A. F. (2022). *Introduction to mediation, moderation, and conditional process analysis: A regression-based approach* (3rd ed.). The Guilford Press.

Henze, N., & Zirkler, B. (1990). A class of invariant consistent tests for multivariate normality. *Communications in Statistics - Theory and Methods*, *19*(10), 3595–3617. https://doi.org/10.1080/03610929008830400

Kessler, R. C., Adler, L., Ames, M., Demler, O., Faraone, S., Hiripi, E., Howes, M. J., Jin, R., Secnik, K., Spencer, T., Ustun, T. B., & Walters, E. E. (2005). The World Health Organization adult ADHD self-report scale (ASRS): A short screening scale for use in the general population. *Psychological Medicine*, *24*(2), 245–256. https://doi.org/10.1017/S0033291704002892

Korkmaz, S., Goksuluk, D., & Zararsiz, G. (2014). MVN: An R package for assessing multivariate normality. *The R Journal*, *6*(2), 151–162. https://doi.org/10.32614/RJ-2014-031

Little, R. J. A. (1988). A Test of Missing Completely at Random for Multivariate Data with Missing Values. *Journal of the American Statistical Association*, *83*(404), 1198–1202. https://doi.org/10.1080/01621459.1988.10478722

Nahhas, R. W. (2024). *Introduction to Regression Methods for Public Health Using R* (1st ed.). Chapman and Hall/CRC. https://doi.org/10.1201/9781003263197

R Core Team. (2025). *R: A language and environment for statistical computing* (Version 4.5.2) [Computer software]. R Foundation for Statistical Computing. https://www.R-project.org/

Rosseel, Y. (2012). lavaan: An R package for structural equation modeling: 1–36. *Journal of Statistical Software*, *48*(2). https://doi.org/10.18637/jss.v048.i02

Rosseel, Y., Jorgensen, T. D., & De Wilde, L. (2025). *lavaan: Latent variable analysis* (Version 0.6-20) [Computer software]. https://doi.org/10.32614/CRAN.package.lavaan

Spitzer, R. L., Kroenke, K., Williams, J. B. W., & Löwe, B. (2006). A brief measure for assessing generalized anxiety disorder: The GAD-7. *Archives of Internal Medicine*, *166*(10), 1092–1097. https://doi.org/10.1001/archinte.166.10.1092

Tierney, N., & Cook, D. (2023). Expanding tidy data principles to facilitate missing data exploration, visualization and assessment of imputations. *Journal of Statistical Software*, *105*(7). https://doi.org/10.18637/jss.v105.i07

Van Rooij, A. J., Schoenmakers, T. M., Van Den Eijnden, R. J. J. M., Vermulst, A. A., & Van De Mheen, D. (2012). Video Game Addiction Test: Validity and psychometric characteristics. *Cyberpsychology, Behavior, and Social Networking*, *15*(9), 507–511. https://doi.org/10.1089/cyber.2012.0007

Young, K. S. (1998). *Caught in the net: How to recognize the signs of Internet addiction--and a winning strategy for recovery*. J. Wiley.
